# COVID-19 infections among Healthcare Workers and Transmission within Households

**DOI:** 10.1101/2020.06.12.20129619

**Authors:** Kevin L. Schwartz, Camille Achonu, Sarah A. Buchan, Kevin A. Brown, Brenda Lee, Michael Whelan, Julie HC Wu, Gary Garber

## Abstract

**Importance:** Protecting healthcare workers (HCWs) from COVID-19 is a priority to maintain a safe and functioning healthcare system. The risk of transmitting COVID-19 to family members is a source of stress for many.

**Objective:** To describe and compare HCW and non-HCW COVID-19 cases in Ontario, Canada, as well as the frequency of COVID-19 among HCWs’ household members.

**Design, Setting, and Participants:** Using reportable disease data at Public Health Ontario which captures all COVID-19 cases in Ontario, Canada, we conducted a population-based cross-sectional study comparing demographic, exposure, and clinical variables between HCWs and non-HCWs with COVID-19 as of 14 May 2020. We calculated rates of infections over time and determined the frequency of within household transmissions using natural language processing based on residential address.

**Exposures and Outcomes:** We contrasted age, gender, comorbidities, clinical presentation (including asymptomatic and presymptomatic), exposure histories including nosocomial transmission, and clinical outcomes between HCWs and non-HCWs with confirmed COVID-19.

**Results:** There were 4,230 (17.5%) HCW COVID-19 cases in Ontario, of whom 20.2% were nurses, 2.3% were physicians, and the remaining 77.4% other specialties. HCWs were more likely to be between 30-60 years of age and female. HCWs were more likely to present asymptomatically (8.1% versus 7.0%, p=0.010) or with atypical symptoms (17.8% versus 10.5%, p<0.001). The mortality among HCWs was 0.2% compared to 10.5% of non-HCWs. HCWs commonly had exposures to a confirmed case or outbreak (74.1%), however only 3.1% were confirmed to be nosocomial. The rate of new infections was 5.5 times higher in HCWs than non-HCWs, but mirrored the epidemic curve. We identified 391 (9.8%) probable secondary household transmissions and 143 (3.6%) acquisitions. Children < 19 years comprised 14.6% of secondary cases compared to only 4.2% of the primary cases.

**Conclusions and Relevance:** HCWs represent a disproportionate number of COVID-19 cases in Ontario but with low confirmed numbers of nosocomial transmission. The data support substantial testing bias and under-ascertainment of general population cases. Protecting HCWs through appropriate personal protective equipment and physical distancing from colleagues is paramount.

**Key Points:** *Question:* What are the differences between healthcare workers and non-healthcare workers with COVID-19?

*Findings:* In this population-based cross-sectional study there were 4,230 healthcare workers comprising 17.5% of COVID-19 cases. Healthcare workers were diagnosed with COVID-19 at a rate 5.5 times higher than the general population with 0.8% of all healthcare workers, compared to 0.1% of non-healthcare workers.

*Meaning:* High healthcare worker COVID-19 burden highlights the importance of physical distancing from colleagues, appropriate personal protective equipment, as well as likely substantial testing bias and under-ascertainment of COVID-19 in the general population.

## Introduction

We are in the midst of a global pandemic from Coronavirus disease of 2019 (COVID-19), caused by Severe Acute Respiratory Syndrome Coronavirus-2 (SARS-CoV-2). COVID-19 is impacting healthcare systems globally which are coping with outbreaks in congregate living facilities and the rapid influx of critically ill patients requiring care in intensive care units.^1^ Preventing healthcare worker (HCW) infections is critical to maintaining a functioning healthcare system, and they have been a priority group for testing throughout the pandemic.^2^ The COVID-19 pandemic has been associated with substantial stress and adverse mental health outcomes for HCWs.^3^

The proportion of COVID-19 cases affecting HCWs from single centre reports has ranged from 0 to 29%.^4-6^ In China approximately 4% of all COVID-19 cases were in HCWs, with an infection rate three times higher in HCWs compared to the general population.^7-9^ HCWs may be exposed to COVID-19 in the community, at work from patients as well as fellow HCWs, and may pose a risk to others around them if infected. The World Health Organization (WHO) has identified research priorities related to the burden and risk factors for HCW COVID-19 infections as well as risk factors for household transmission from HCWs.^3^ To our knowledge no studies have evaluated transmission of COVID-19 from HCWs to household contacts. Our objectives were to describe demographic, exposure, and clinical symptom differences between HCW and non-HCW COVID-19 cases in Ontario, as well as the frequency of HCW household members with COVID-19.

## Methods

### Design and Setting

We conducted a cross-sectional study comparing HCW and non-HCW COVID-19 cases. Data collection began with the first COVID-19 diagnosed patient in Ontario, Canada on 21 January 2020 until 14 May 2020. We obtained the data from reportable surveillance infectious disease data at Public Health Ontario (PHO). The activities described in this manuscript were conducted in fulfillment of PHO’s legislated mandate to provide scientific and technical advice and operational support in an emergency or outbreak situation *(<u>Ontario Agency for Health Protection and Promotion Act, SO 2007, c 10</u>*). Research ethics committee approval was sought and determined to be not required because the activities described are considered public health practice and not research.

### Data Source

We obtained the data from the integrated Public Health Information System (iPHIS), the Toronto Public Health Coronavirus Rapid Entry System (CORES), the Ottawa Public Health COVID-19 Ottawa Database (The COD), and Middlesex-London COVID-19 Case and Contact Management tool (CCMtool) accessed on 25 May 2020 (but only including cases up to 14 May 2020 to account for a delay in reporting). These databases are web-based information systems for the reporting and surveillance of diseases of public health significance in Ontario. PHO is a Crown corporation dedicated to protecting and promoting the health of all Ontarians and reducing inequities in health. All probable or confirmed COVID-19 positive individuals are entered by local public health units.

### Definitions and Variables

A HCW was defined as an individual with an occupation involving caring for patients. All other individuals were classified as non-HCWs. We used confirmed COVID-19 cases only. Demographic information available included gender, age, comorbidities (anemia, asthma, cancer, cardiovascular condition, chronic liver disease, chronic obstructive pulmonary disease, diabetes, immunocompromised, neurologic disorder, obesity, pregnancy or 6 weeks post-partum, renal condition, tuberculosis, and other chronic medical condition). Exposures were classified by the local public health unit contact investigation as outbreak associated or close contact to a confirmed or probable case of COVID-19, community transmission with no epidemiological link, travel to an endemic area for COVID-19 outside of Ontario within the incubation period of 14 days, or missing exposure information. A separate variable to identify nosocomial cases was added 3 April 2020. Analysis with the nosocomial variable was limited to this time frame. Clinical symptoms were classified as asymptomatic, presymptomatic (defined as having a testing date prior to symptoms onset date), typical with fever and/or cough, atypical (any other symptom), or missing. Clinical outcomes were classified in descending order as died, requiring a ventilator, intensive care unit without a ventilator, hospitalized, or not hospitalized.

Onset of illness was defined as symptom onset date, which was available for 66.3% of the cohort. For those missing symptom onset date we calculated the median number of days from test date to symptom onset date where the data was available (median=4 days) and performed a deterministic imputation. If test date was not available we used the median time from symptoms onset to date reported to the local public health unit (median=5 days). Onset date in asymptomatic individuals was the testing date or reporting date if not available. We defined household spread using a natural language processing algorithm to link confirmed HCW COVID-19 cases to other probable or confirmed COVID-19 cases by residential address. If symptom onset dates in the non-HCWs were two or more days earlier than the HCWs then these were defined as a probable HCW *acquisition*. If household cases symptom onset dates were two or more days following a HCWs’ then this was defined as a probable *transmission*. Infections that were -1, 0, or +1 days apart were classified as unknown direction of transmission. As a sensitivity analysis we used ±4 days for greater confidence in the direction of transmission.

### Statistical Analysis

Variables were compared between HCWs and non-HCWs by chi-squared tests, t-tests, or non-parametric tests as appropriate in bivariate analyses with two-side p-values <0.05 as statistically significant. Rates of infection in non-HCWs were determined using population denominators from Statistics Canada. HCW denominators were calculated from publically available sources at the Canadian Institute for Health Information and Statistics Canada.^10,11^

## Results

There are an estimated 552,560 HCWs and 14,311,868 non-HCWs in Ontario. As of 14 May 2020 there were 24,202 confirmed COVID-19 cases in Ontario, including 4,230 (17.5%) HCWs. There was geographical variability in the proportion of COVID-19 HCW cases ranging from 4-42% across Ontario’s 34 public health units. In general, the regions with the largest numbers of cases did not overlap with those with the highest proportions (Figure 1). For instance, Toronto and Peel regions had the highest numbers of cases (1,315 and 403), but were in the lower half of regions by the proportion of cases that were HCWs (15.3% and 11.2%), at the time of this analysis.

**Figure 1:**
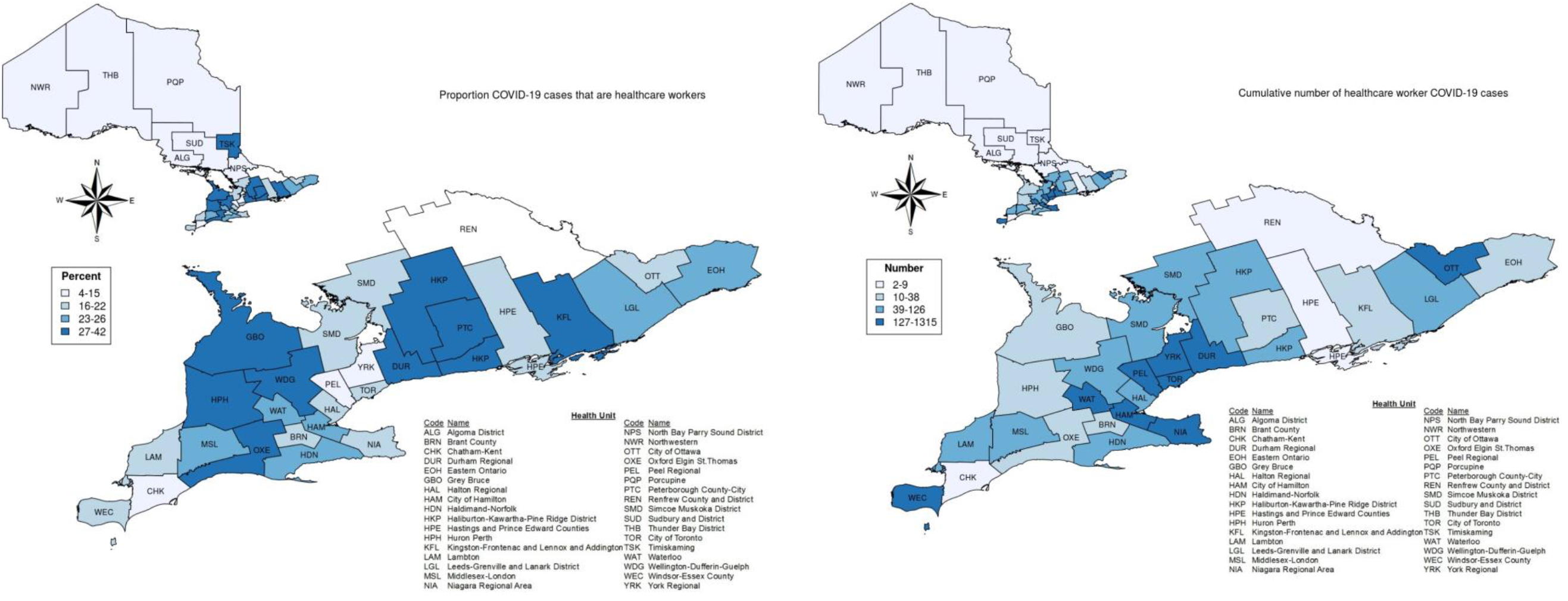
Geographical variability by Ontario public health unit. Left map shows the percent of total COVID-19 cases in each region that are healthcare workers. Right map shows the cumulative number of healthcare worker COVID-19 cases.

HCWs with COVID-19 were more likely to be female and were more commonly between the ages of 30-60 years compared to non-HCWs. There were 4 (0.1%) HCWs ≥75 years of age compared to 6,158 (30.8%) non-HCWs. Approximately one quarter of both HCW and non-HCWs had one or more comorbidities (Table 1).

**Table 1:**
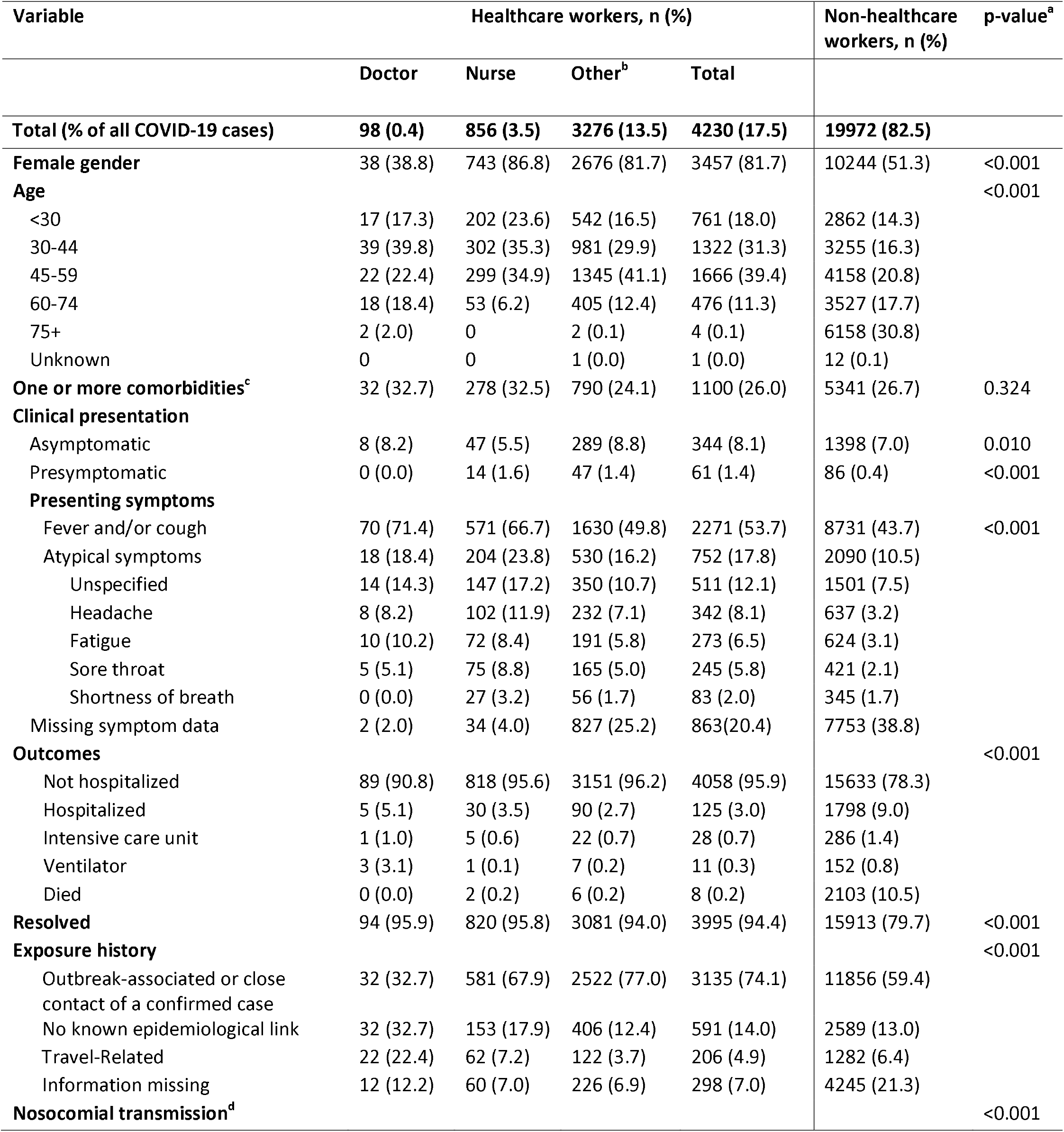
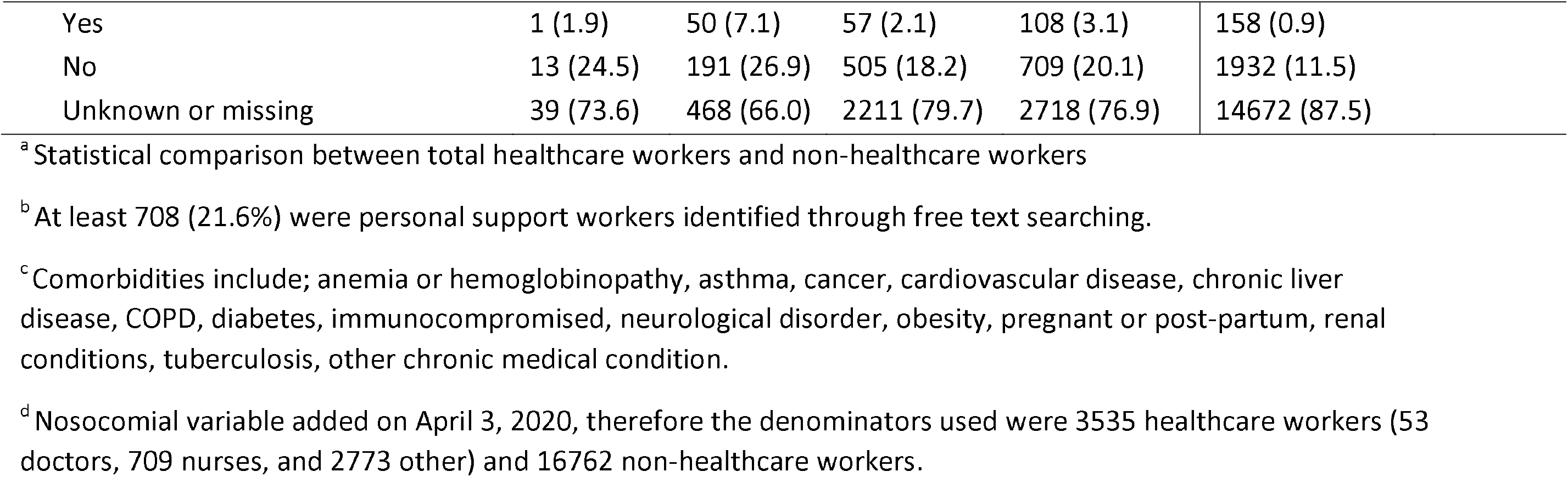
Demographic and exposure comparison between healthcare worker and non-healthcare worker COVID-19 cases (n=24,202).

| Variable | Healthcare workers, n (%) |  |  |  | Non-healthcare workers, n (%) | p-value<sup>a</sup> |
| --- | --- | --- | --- | --- | --- | --- |
|  | Doctor | Nurse | Other<sup>b</sup> | Total |  |  |
| **Total (% of all COVID-19 cases)** | **98 (0.4)** | **856 (3.5)** | **3276 (13.5)** | **4230 (17.5)** | **19972 (82.5)** |  |
| **Female gender** | 38 (38.8) | 743 (86.8) | 2676 (81.7) | 3457 (81.7) | 10244 (51.3) | <0.001 |
| **Age** |  |  |  |  |  | <0.001 |
| <30 | 17 (17.3) | 202 (23.6) | 542 (16.5) | 761 (18.0) | 2862 (14.3) |  |
| 30-44 | 39 (39.8) | 302 (35.3) | 981 (29.9) | 1322 (31.3) | 3255 (16.3) |  |
| 45-59 | 22 (22.4) | 299 (34.9) | 1345 (41.1) | 1666 (39.4) | 4158 (20.8) |  |
| 60-74 | 18 (18.4) | 53 (6.2) | 405 (12.4) | 476 (11.3) | 3527 (17.7) |  |
| 75+ | 2 (2.0) | 0 | 2 (0.1) | 4 (0.1) | 6158 (30.8) |  |
| Unknown | 0 | 0 | 1 (0.0) | 1 (0.0) | 12 (0.1) |  |
| **One or more comorbidities<sup>c</sup>** | 32 (32.7) | 278 (32.5) | 790 (24.1) | 1100 (26.0) | 5341 (26.7) | 0.324 |
| **Clinical presentation** |  |  |  |  |  |  |
| Asymptomatic | 8 (8.2) | 47 (5.5) | 289 (8.8) | 344 (8.1) | 1398 (7.0) | 0.010 |
| Presymptomatic | 0 (0.0) | 14 (1.6) | 47 (1.4) | 61 (1.4) | 86 (0.4) | <0.001 |
| **Presenting symptoms** |  |  |  |  |  |  |
| Fever and/or cough | 70 (71.4) | 571 (66.7) | 1630 (49.8) | 2271 (53.7) | 8731 (43.7) | <0.001 |
| Atypical symptoms | 18 (18.4) | 204 (23.8) | 530 (16.2) | 752 (17.8) | 2090 (10.5) |  |
| Unspecified | 14 (14.3) | 147 (17.2) | 350 (10.7) | 511 (12.1) | 1501 (7.5) |  |
| Headache | 8 (8.2) | 102 (11.9) | 232 (7.1) | 342 (8.1) | 637 (3.2) |  |
| Fatigue | 10 (10.2) | 72 (8.4) | 191 (5.8) | 273 (6.5) | 624 (3.1) |  |
| Sore throat | 5 (5.1) | 75 (8.8) | 165 (5.0) | 245 (5.8) | 421 (2.1) |  |
| Shortness of breath | 0 (0.0) | 27 (3.2) | 56 (1.7) | 83 (2.0) | 345 (1.7) |  |
| Missing symptom data | 2 (2.0) | 34 (4.0) | 827 (25.2) | 863 (20.4) | 7753 (38.8) |  |
| **Outcomes** |  |  |  |  |  | <0.001 |
| Not hospitalized | 89 (90.8) | 818 (95.6) | 3151 (96.2) | 4058 (95.9) | 15633 (78.3) |  |
| Hospitalized | 5 (5.1) | 30 (3.5) | 90 (2.7) | 125 (3.0) | 1798 (9.0) |  |
| Intensive care unit | 1 (1.0) | 5 (0.6) | 22 (0.7) | 28 (0.7) | 286 (1.4) |  |
| Ventilator | 3 (3.1) | 1 (0.1) | 7 (0.2) | 11 (0.3) | 152 (0.8) |  |
| Died | 0 (0.0) | 2 (0.2) | 6 (0.2) | 8 (0.2) | 2103 (10.5) |  |
| **Resolved** | 94 (95.9) | 820 (95.8) | 3081 (94.0) | 3995 (94.4) | 15913 (79.7) | <0.001 |
| **Exposure history** |  |  |  |  |  | <0.001 |
| Outbreak-associated or close contact of a confirmed case | 32 (32.7) | 581 (67.9) | 2522 (77.0) | 3135 (74.1) | 11856 (59.4) |  |
| No known epidemiological link | 32 (32.7) | 153 (17.9) | 406 (12.4) | 591 (14.0) | 2589 (13.0) |  |
| Travel-Related | 22 (22.4) | 62 (7.2) | 122 (3.7) | 206 (4.9) | 1282 (6.4) |  |
| Information missing | 12 (12.2) | 60 (7.0) | 226 (6.9) | 298 (7.0) | 4245 (21.3) |  |
| **Nosocomial transmission<sup>d</sup>** |  |  |  |  |  | <0.001 |

|  |  |  |  |  |  |
| --- | --- | --- | --- | --- | --- |
| Yes | 1 (1.9) | 50 (7.1) | 57 (2.1) | 108 (3.1) | 158 (0.9) |
| No | 13 (24.5) | 191 (26.9) | 505 (18.2) | 709 (20.1) | 1932 (11.5) |
| Unknown or missing | 39 (73.6) | 468 (66.0) | 2211 (79.7) | 2718 (76.9) | 14672 (87.5) |

<sup>a</sup> Statistical comparison between total healthcare workers and non-healthcare workers
<sup>b</sup> At least 708 (21.6%) were personal support workers identified through free text searching.
<sup>c</sup> Comorbidities include; anemia or hemoglobinopathy, asthma, cancer, cardiovascular disease, chronic liver disease, COPD, diabetes, immunocompromised, neurological disorder, obesity, pregnant or post-partum, renal conditions, tuberculosis, other chronic medical condition.
<sup>d</sup> Nosocomial variable added on April 3, 2020, therefore the denominators used were 3535 healthcare workers (53 doctors, 709 nurses, and 2773 other) and 16762 non-healthcare workers.

Asymptomatic infections occurred in 8.1% of HCWs and 7.0% of non-HCWs (p=0.010). Presymptomatic infections were infrequently identified but more common in HCWs (1.4% versus 0.4%, p<0.001). Of those with symptoms at diagnosis, atypical symptoms were more common in HCWs (17.8% versus 10.5%, p<0.001). The most common atypical symptoms were headache, fatigue, and sore throat (Table 1). HCWs had milder clinical courses represented by 95.9% not requiring hospitalization and only 8 (0.2%) deaths, compared to non-HCWs with 78.3% managed outside of hospital with 2,103 (10.5%) deaths (p<0.001).

HCWs were identified as cases at a rate 5.5 times higher than non-HCWs with rates of 765.5 per 100,000 compared to 139.5 per 100,000. There were 98 physicians infected, comprising 2.3% of HCW infections, with an infection rate approximately two times the general population. Nurses comprised 20.2% of HCW infections and 3.5% of all Ontario infections with an infection rate of approximately four times higher than the general population. Other HCWs, which includes personal support workers (PSWs), comprised the largest group with an infection rate of 6.5 times that of the general population (Table 2).

**Table 2:**
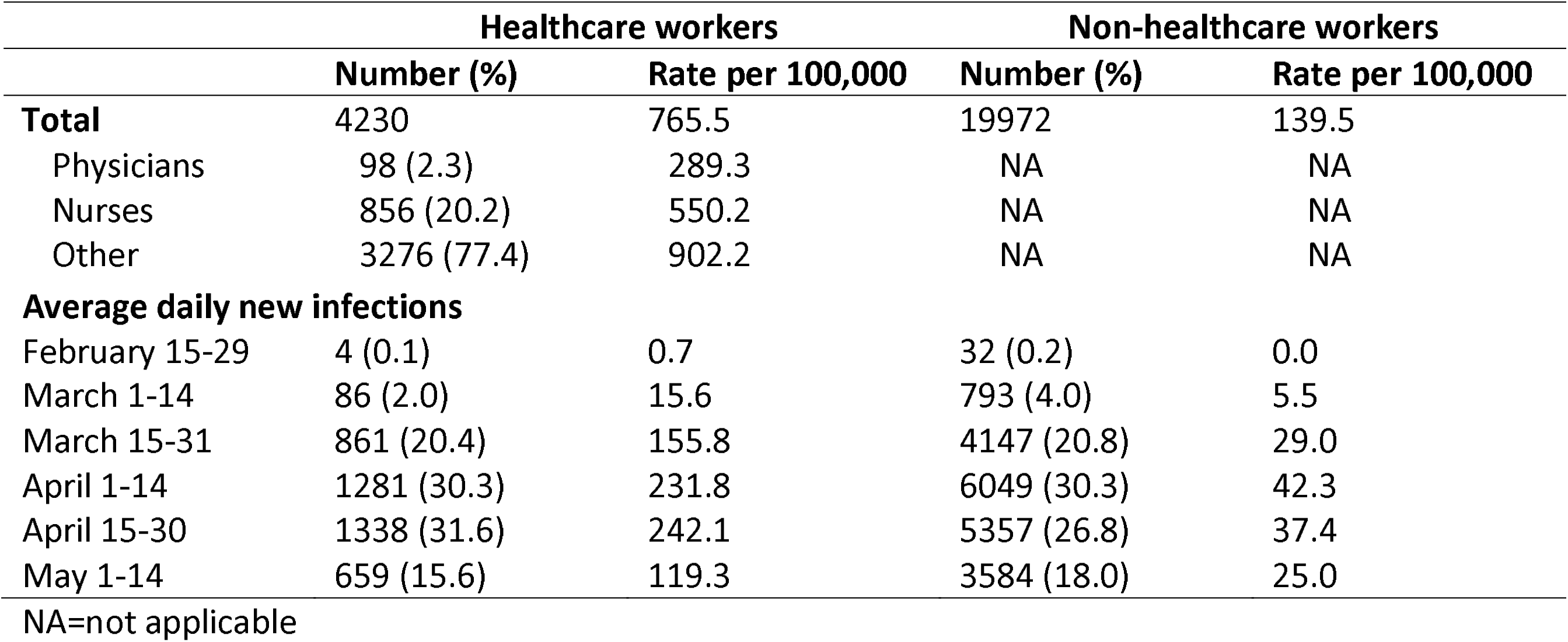
Rate of new COVID-19 cases in healthcare workers and non-healthcare workers

The difference in daily new infection rates varied between 2-6 fold throughout the epidemic, mirroring the epidemic curve (Figure 2). As testing capacity improved in Ontario the difference in detection rates began to fall at the end of April and early May.

**Figure 2:**
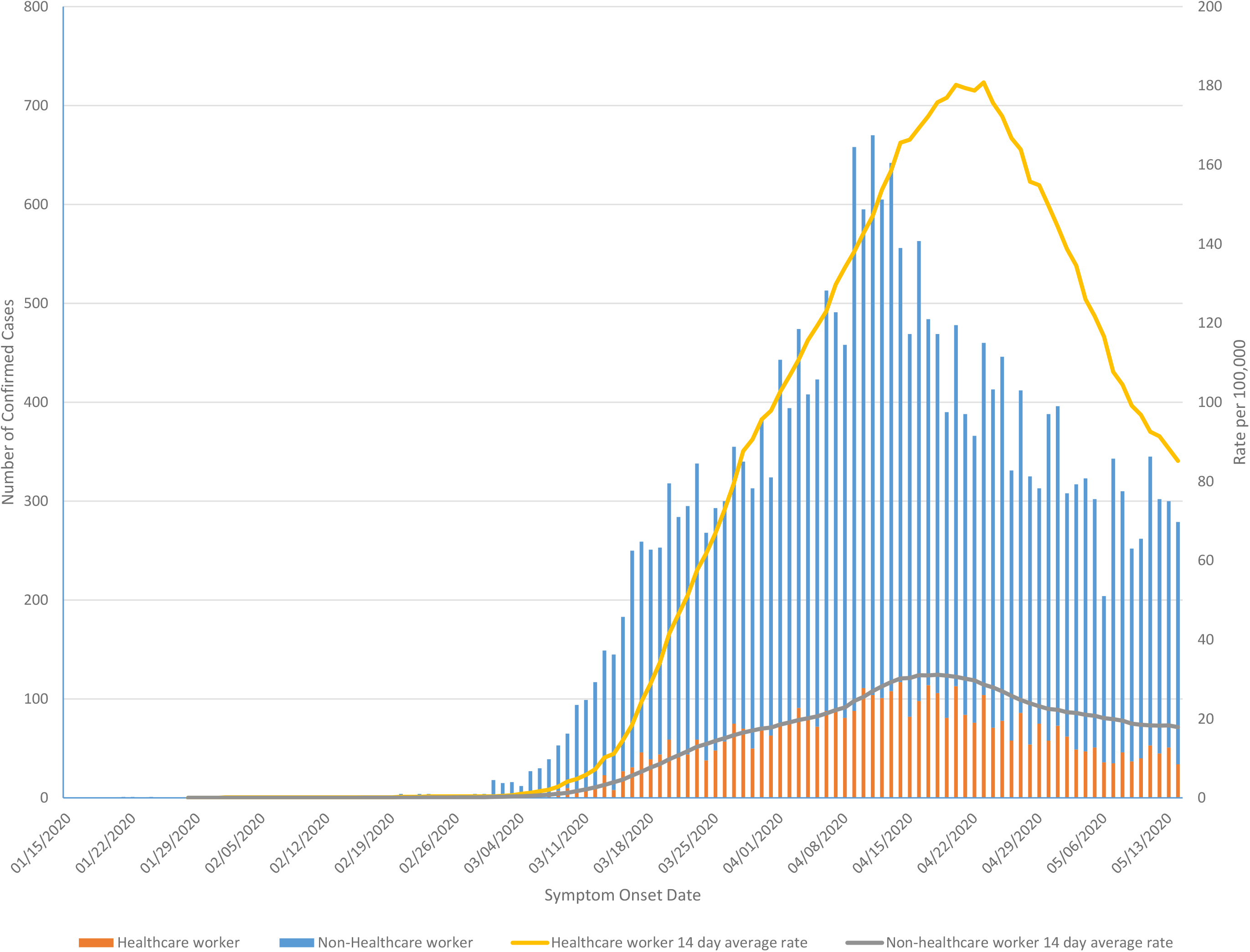
Epidemic curve by symptom onset date showing daily new case numbers (left axis) and 14-day moving average of daily rate of new COVID-19 cases (right axis) for healthcare workers and non-healthcare workers.

From the available exposure history data, 74.1% of HCWs either had contact with a confirmed cases or were associated with an outbreak. A recent travel history was overall uncommon (4.9%), except for doctors (22.4%). Only 3.1% of HCWs were documented to have acquired their infection nosocomially in the subset of HCWs with this data available. Nurses had the highest risk of nosocomial transmission (7.1%) (Table 1).

There were 675 (16.9%) HCW household cases, with a median of 1 and a range of 1-5 household cases. We observed that 391 (9.8%) of HCWs probably transmitted COVID-19 to a household member; of these, 14.6% were to children <19 years, 65.7% to those 19-59 years and 19.7% to those ≥60 years. We observed 143 (3.6%) instances where the HCW probably acquired the infection from a household contact; of these, 4.2% were from children <19 years, 62.9% from adults 19-59 years and 32.9% from adults ≥60 years. In 141 (3.5%) contacts the direction of transmission could not be determined (Table 3). In the sensitivity analysis using ±4 days, instead of ±2 days, there were 275 (6.9%) transmissions and 90 (2.3%) acquisitions.

**Table 3:** Healthcare worker within household COVID-19 transmissions (n=3986 healthcare workers)

|  | Number (%) | Age group of household contact, n (row %) |  |  |
| --- | --- | --- | --- | --- |
|  |  | <19 years | 19-59 years | ≥60 years |
| **HCW acquired from household contact** | 143 (3.6) | 6 (4.2) | 90 (62.9) | 47 (32.9) |
| **HCW transmitted to household contact** | 391 (9.8) | 57 (14.6) | 257 (65.7) | 77 (19.7) |
| **Household infection with unknown direction of transmission** | 141 (3.5) | 19 (13.5) | 94 (66.7) | 28 (19.9) |
| **Total household transmissions involving HCWs** | 675 (16.9) | 82 (12.1) | 441 (65.3) | 152 (22.5) |

HCW=healthcare worker; 244 HCWs did not have a home address listed

## Interpretation

HCWs have comprised 17.5% of the 24,202 confirmed COVID-19 cases in Ontario as of May 14, 2020. The rate of new infections per day varied between two and six times the general population over time and by type of HCW with physicians being lower risk than nurses, who were lower risk than other specialties combined, which includes PSWs.

Data from China has observed that approximately 4% of all COVID-19 cases were in HCWs.^8,9^ There has been wide variation in single centre reports of HCW COVID-19 infections. Early in the pandemic one report from Wuhan observed that 29% of 138 hospitalized COVID-19 cases were in HCWs, with at least 10 of those related to a possible super-spreader event.^6^ However, other centres have reported no COVID-19 cases despite significant exposures.^4,12^ In a long term care facility outbreak in Washington state, HCWs comprised 26% of infections, however the clinical course was substantially less severe in HCWs compared to residents and visitors.^5^ The Center for Disease Control and Prevention in the United States reported on over 9,000 infected HCWs. Compared to our study they observed higher rates of hospitalizations (10% versus 3% in our study) but similar mortality (0.3% versus 0.2%).^13^

Studies from China and the Netherlands reported that approximately 1% of HCWs within hospitals were infected with COVID-19.^14,15^ In our study, 0.8% of all HCWs and 0.1% of non-HCWs in Ontario have been identified as COVID-19 cases, including 0.3% of doctors and 0.6% of nurses. A report from Alberta, Canada identified that 0.1% of all HCWs were COVID-19 cases (including 0.3% of doctors) compared to 0.1% of the general population.^16^ Alberta has similar IPAC guidance on personal protective equipment (PPE) to Ontario which includes surgical masks, eye protection, gloves, and gowns for suspected or confirmed COVID-19 patients and N95 respirators for aerosol generating medical procedures. Our study was not able to evaluate the adequacy of PPE used by HCWs, however it is unlikely to explain the differences between Ontario and Alberta. There are multiple potential explanations for the higher rate in Ontario HCWs. Ontario likely has substantially more undiagnosed cases in the general population as well as more long-term care home outbreaks. However, we were unable to determine from our data which HCWs worked in long-term care. We could not reconcile the large number of outbreak associated cases with the low numbers of household acquisition (2-4%) and nosocomial acquisition (3.1%). One explanation is that a single long term care home case in either a HCW or resident did qualify as an “outbreak” in Ontario. It is possible that HCWs are acquiring COVID-19 at work from fellow HCWs or asymptomatic patients. In Alberta only 12 (8.8%) HCW COVID-19 cases were from occupational exposures.^16^ Further study is needed to better understand the nosocomial risk of COVID-19 among HCWs.

A study from the United Kingdom prospectively tested HCWs at a large hospital and found that while positivity was high it did not vary between patient-facing HCWs (15%), non-clinical HCWs with nosocomial risk factors (16%), and low risk HCWs without nosocomial risk (18%).^17^ A study from Wuhan, China observed a cumulative incidence of COVID-19 in 1.1% (110 of 9685) of HCWs. However, the rates were 0.5% among HCWs with direct contact to suspected or confirmed COVID-19 patients, 1.6% in HCWs with patient contact in non-COVID departments, and 1.0% among HCWs with no patient contact.^14^ The higher risk of infection in both non-COVID-19 clinical areas and non-clinical areas, suggests that HCWs may be acquiring COVID-19 from co-workers or in the community. We feel the data highlight the importance of maintaining physical distancing from colleagues (i.e. in break room or during meals) or utilizing additional PPE when physical distancing cannot be achieved. A policy for universal masking of HCWs was recently adopted in Ontario, and the importance of this policy in protecting both patients as well as fellow HCWs should be emphasized.

Testing bias is almost certainly contributing to the discrepancy in HCW and non-HCW rates. In a report from Lombardy, Italy that performed serological studies on HCWs and non-HCWs, they identified that 23% of HCWs sampled were positive for COVID-19 antibodies compared to 62% of the general population.^18^ Furthermore, we identified dramatically different mortality rates between HCWs and non-HCWs. This is likely due to both age differences and substantial underestimates of true population disease burden, which may actually be closer to the rate we observed in HCWs. HCWs have been a priority population for testing throughout the pandemic, even during times of limited capacity. Until the last two weeks of the study period COVID-19 testing was generally restricted to high risk groups (i.e. hospitalized patients, HCWs, other essential workers, vulnerable populations). Population based serological studies will enable a better understanding of this discrepancy.

HCWs may be a potential source of COVID-19 transmission to both patients and household contacts. In particular since it has become apparent that presymptomatic cases comprise a substantial portion of transmissions.^19^ Transmitting to household members has been a source of stress for HCWs^20^ and has been identified as a knowledge gap by the WHO.^3^ To our knowledge this is the first study evaluating within household transmission risks associated with HCWs. We observed that 9.8% of infected HCWs likely transmitted COVID-19 to a household contact. Using a stricter definition of 4 days, we observed a transmission rate of 6.9%. Interestingly, children were infrequently the source of infection compared to secondary household cases (4.2% versus 14.6%), consistent with prior research demonstrating that children may be inefficient transmitters of COVID-19.^21^

This study has some limitations. The data quality is dependent on entry by 34 public health units across Ontario. Data completeness and quality may vary, and it is possible some HCWs were misclassified. The variable for nosocomial transmission was added later in the study period and largely incomplete even after restricting to the later time period. This may be in part due to the challenge of assigning causality of infections without a clear exposure history or multiple potential exposures. The general population, doctor, and nurse denominators used to calculate infection rates are accurate and current, however there is no complete data source for other HCWs in Ontario. We used various sources to arrive at an overall estimated HCW denominator which may impact the ability to compare rates. We lacked granularity in the type of HCW beyond physician or nurse (i.e. PSW) as well as the setting of employment (e.g. long-term care, hospital, etc.). Finally, while HCWs were priority groups for testing throughout the pandemic, testing criteria and capacity varied substantially. Case identification was substantially better throughout for HCWs and as a result general population cases are undoubtedly underestimates.

In conclusion, 0.8% of HCWs in Ontario have been diagnosed with COVID-19. We identified clinical, demographic, and geographical differences from over 4,000 HCWs, compared to almost 20,000 non-HCWs. Despite these large numbers, relatively few were linked to a nosocomial exposure. Further system improvements and monitoring are needed to protect all HCWs from COVID-19 with an emphasis on physical distancing from colleagues and appropriate use of PPE.

## Data Availability

Data is not publically available

## Acknowledgement

We would like to thank James Johnson for developing the natural language processing algorithm linking household cases, as well as Brendan Smith and Christine Warren for their help determining total numbers of healthcare workers in Ontario.

